# Using geospatial models to map zero-dose children: factors associated with zero-dose vaccination status before and after a mass measles and rubella vaccination campaign

**DOI:** 10.1101/2021.09.16.21263705

**Authors:** Rohan Arambepola, Yangyupei Yang, Kyle Hutchinson, Francis D. Mwansa, Julie Ann Doherty, Frazer Bwalya, Phillimon Ndubani, Gloria Musukwa, William J. Moss, Amy Wesolowski, Simon Mutembo

## Abstract

**Introduction:** Despite gains in global coverage of childhood vaccines, many children remain undervaccinated. Vaccination campaigns also known as Supplemental Immunization Activity (SIA) are commonly conducted to reach those who are undervaccinated. However, reaching these children even during an SIA is challenging. We evaluated the effectiveness of an SIA in reaching zero dose children.

**Methods:** We conducted a prospective study in 10 health center catchment areas in Southern province, Zambia in November 2020. About 2 months before the measles and rubella SIA we developed aerial satellite maps which were then used to enumerate and survey households. Zero dose children were identified during this exercise. After the SIA, households with zero dose children identified before the SIA were targeted for mop up vaccination and to assess if they were vaccinated during the SIA. A Bayesian geospatial model was used to identify factors associated with zero-dose status before the campaign and produce fine-scale prevalence maps. Models were used to identify factors associated with measles zero-dose children reached in the campaign and identify optimal locations for additional vaccination sites.

**Results:** Before the vaccination campaign, 4% of children under 9 months were DTP zero-dose and 17% of children 9-60 months were measles zero-dose. Of the 461 measles zero-dose children identified before the vaccination campaign, 338 (73.3%) were vaccinated during the campaign, 118 (25.6%) were reached by a targeted mop-up activity. The presence of other children in the household, younger age, greater travel time to health facilities, and living between health facility catchment areas were associated with zero-dose status. Mapping zero-dose prevalence revealed substantial heterogeneity, both within and between catchment areas. Several potential locations were identified for additional vaccination sites.

**Conclusion:** Fine-scale variation in zero-dose prevalence and the impact of accessibility to healthcare facilities on vaccination coverage were identified. Geospatial modeling can aid targeted vaccination activities.

**Summary:** *What is already known?:*

- In many low- and lower-middle income countries, improvements in routine childhood vaccination coverage have stalled.
- An estimated 17 million children globally have not received any routine vaccinations (zero-dose children).
- Zero-dose children, and those who have not received any doses of specific vaccines such as measles, remain vulnerable to preventable diseases and can sustain transmission in otherwise highly vaccinated populations
- A lack of understanding of the number and spatial distribution of zero-dose children make targeting vaccination activities to reach this group challenging.

*What are the new findings?:*

- Prior to a mass measles and rubella vaccination campaign, 17% of children younger than 9 months of age in the study area had not received the DTP vaccine and 4% of children 9 months or older had not received a measles-containing vaccine.
- Over a quarter of the children identified as not having received a measles-containing vaccine before the measles and rubella mass vaccination campaign were not vaccinated during the campaign.
- Geospatial models revealed substantial fine-scale variation in zero-dose status and optimal locations for additional vaccination sites.

*What do the new findings imply?:*

- There is potential for using similar household-level geospatial survey and modeling strategies to improve targeting of vaccination activities to reach zero-dose children.

## INTRODUCTION

Significant gains in vaccination coverage have been made globally since the 1980s due to substantial investment in childhood immunization services. However, in the past decade, progress has stagnated and routine vaccination coverage has declined in many countries [1,2]. Widespread disruption to vaccine delivery due to the COVID-19 pandemic has compounded this problem, with global coverage of the first dose of a diphtheria-tetanus-pertussis-containing vaccine (DTP1) and first-dose of a measles-containing vaccine (MCV1) falling from 90% and 86% in 2019 to 87% and 84% in 2020, respectively [3,4]. This stalling progress and disruption to routine vaccination activities has resulted in pockets of unvaccinated and undervaccinated communities. Communities where vaccination rates are below herd immunity thresholds are at risk of outbreaks of vaccine-preventable diseases [5–7].

These communities at risk of outbreaks are made up of children who missed some vaccine doses as well as children who did not receive any routine vaccinations, the latter referred to as zero-dose children. In practice zero-dose children are often defined as children who have not received a DTP1 vaccine [8–10].There were an estimated 17 million zero-dose children in 2020, the majority living in sub-Saharan Africa or conflict-affected areas [4,9,11,12]. Research on the “immunization cascade”, which describes how children move from zero-dose to fully vaccinated, suggests that in many low- and lower-middle income countries vaccine coverage is polarized, with most children either receiving all or almost all vaccines, or few to none [13]. In many countries, including Zambia, mass measles and rubella vaccination campaigns are carried out with the aim of vaccinating children who have not received their routine doses. Understanding the prevalence and spatial distribution of children in the community who have received few vaccines or are zero-dose is challenging, as these children are likely to be less engaged with routine healthcare services [14]. Additionally, it can be difficult to determine if underserved communities were reached after mass vaccination campaigns.

Several studies have investigated risk factors for low vaccine coverage or incomplete vaccination, often identifying accessibility to healthcare, maternal education, parental attitude, and socioeconomic status as important factors [15–20]. However, remote or marginalized communities may be underrepresented in studies that recruit participants from those already engaged with the formal healthcare system, for example, studies requiring child health cards for eligibility [15,16]. Furthermore, there has been limited focus on zero-dose children in particular, even though interventions that improve overall vaccination coverage can have limited impact on zero-dose children [20]. Post-campaign coverage surveys may be used to identify communities not reached or factors impacting campaign coverage, but these surveys have limited spatial and demographic detail and may also suffer from selection bias [21]. Moreover, it is unclear how to operationalize the results of these studies on vaccination status or campaign coverage to improve the effectiveness of vaccination activities. Data and models that can be used to locate communities with a high prevalence of zero-dose children may help programs better allocate existing services. However, data from health facilities or number of doses given in a campaign cannot capture fine-scale spatial heterogeneities in coverage and are unlikely to be representative.

Detailed household mapping of eligible and vaccinated children pre- and post a national campaign can provide more information on the distribution of zero-dose children and which of these children are vaccinated during vaccination campaigns, as well as provide a framework to model the effectiveness of various campaign strategies, including location of vaccine outreach sites. We describe the household enumeration of children eligible for a mass measles and rubella vaccination campaign conducted in Zambia in November 2020 and a targeted mop-up vaccination activity after the campaign. The mop-up focused on children identified as measles zero-dose in the initial household enumeration (children who were eligible for but had not received any dose of a measles-containing vaccine) to identify which measles zero-dose children were vaccinated during the campaign and vaccinate those who were not. We then developed a geospatial model using these data to estimate fine-scale heterogeneity in vaccination status, which may be missed by more spatially aggregated coverage estimates, and predict the effect of adding new vaccination sites in different locations.

## METHODS

### Study design and population

The study was conducted in 10 health facility catchment areas of Choma District, Southern Province, Zambia (Figure 1). Two of the catchment areas (Choma Railway Surgery and Shampande) are predominantly densely populated urban settings. The remaining catchment areas are rural areas populated by subsistence farmers living in scattered homesteads, characteristic of much of rural sub-Saharan Africa [22]. Overall, reported vaccine coverage in Choma District is high (coverage for DTP1= 99%, MCV1 = 93% and MCV2 = 73% in 2020 [23].

**Figure 1:**
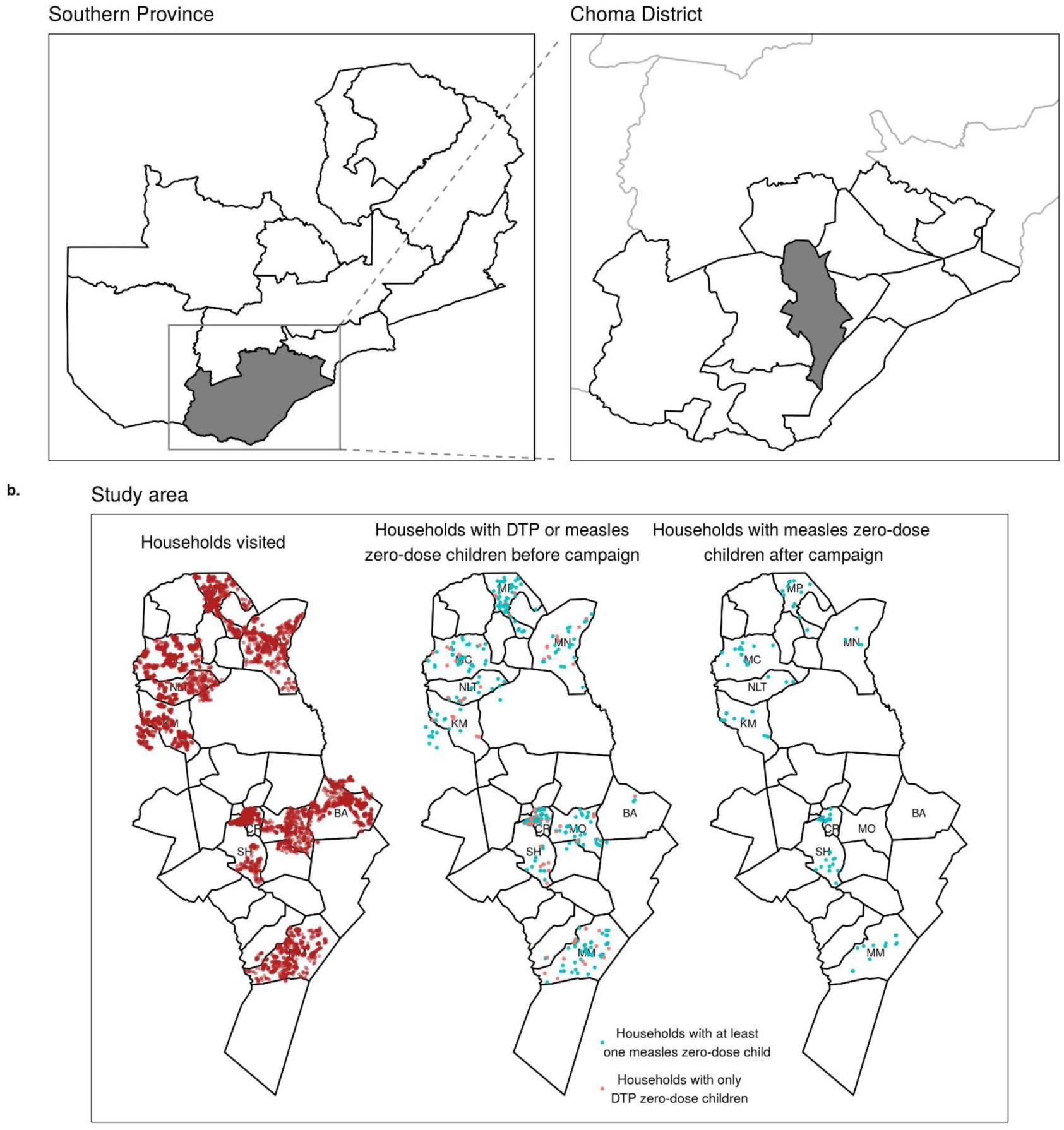
Study area. (a) Location of Choma District within Zambia and Southern Province. (b) Health facility catchment areas in Choma District with the 10 catchment areas in this study labelled: Batoka (BA), Choma Railway Surgery (CR), Kamwanu (KM), Macha (MC), Masuku Mission (MM), Mangunza (MN), Mpanza (MP), Nalituba (NLT), Shampande (SH). The points represent the locations of all households visited (left), households containing zero-dose children before the campaign (middle, shaded in blue for households with any measles zero-dose children) and households containing measles zero-dose children that were not vaccinated during the campaign (right).

We conducted a prospective study to quantify the vaccination status of children before and after a mass measles and rubella vaccination campaign conducted between 20 - 29th November 2020. Approximately 6 weeks prior to the campaign, all structures identified in the study area using satellite imagery were visited by community health volunteers (CHVs) and eligible children were registered. Children between 9 and 60 months of age were eligible to participate, to match eligibility for the mass vaccination campaign. Data were also collected for children below 9 months to gather information on DTP1 vaccination status. These data included age, sex, and measles and rubella vaccination status for children 9 months and older or DTP1 vaccination status for children younger than 9 months. For the purposes of this study, a child was defined as DPT zero-dose if they were younger than 9 months of age and had not received DTP1 and measles zero-dose if they were 9 months or older and had not received any dose of MCV. We did not collect information on DTP1 for children over 9 months. Here, zero-dose status refers to both groups. One week after the vaccination campaign, households where measles zero-dose children were identified were revisited to see if these children were vaccinated during the campaign. If the child was not vaccinated, a measles and rubella vaccine dose was provided.

### Data collection

Data were collected on electronic tablets using Reveal, an open-source platform for mapping populations and monitoring coverage of health interventions by household [24]. Manual and automated enumeration methods and algorithms were applied to high resolution satellite imagery to identify structures within the 10 health facility catchment areas. The operational health facility catchment area boundaries and major landmarks were mapped in a participatory manner with health facility staff and community health workers using google earth. These data were uploaded to the mobile component of the Reveal platform in the form of digital base-maps that assisted field teams with real-time navigation capabilities, to ensure all structures within a given area were identified and reached. Teams of CHVs, using Reveal’s mobile, map-based interface, then went door-to-door to obtain informed consent and register children eligible for the upcoming vaccination campaign. After the campaign, teams of CHVs and health center nurses used the Reveal interface to navigate back to the houses where measles zero-dose children were identified before the campaign to verify whether these children were vaccinated during the campaign.

### Data analysis

#### Univariate analyses

The relationships between several factors and zero-dose status before the mass vaccination campaign were initially assessed in univariate analyses. These factors were age, travel time to the nearest health facility calculated using a friction surface [25] (see Supplementary material section 2 for more details), the presence of another eligible child in the household, and whether the household was “between health facilities”. A household was defined as being between health facilities if travel time to the second closest health facility was within 20% of the travel time to the nearest health facility. The location of these between facility households is shown in Supplementary Figure S1 and this percentage was varied in a sensitivity analysis also detailed in Supplementary material section 3. The relationship between the likelihood of a measles zero-dose child being vaccinated in the campaign and travel time to the nearest vaccination site was also investigated.

#### Geostatistical model

Separate Bayesian geospatial models, based on the geostatistics framework pioneered by Diggle and others [26–28], were used to model DTP zero-dose and measles zero-dose prevalence across the study area. Let *y*_*i*_ be the missing vaccination status of child 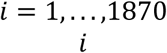 for DTP and *i* = 1, …, 11649 for measles), that is *y*_*i*_ was 1 if the child had not received DTP1 (if under 9 months) or MCV1 (if 9 months and older) and 0 if they had. Let *l*_*i*_ be the location of the household this child lived in. Missing vaccination status was modelled as a realization of a Bernoulli process,

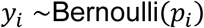

with underlying probability *p*_*i*_for a child of this age at this location. This underlying probability was on a logit-scale as the sum of a linear contribution from covariates and a Gaussian process over space

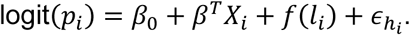

Here *X*_*i*_ were covariate values, *f* was the Gaussian process term and *β*_0_, *β* were parameters to be learned. The covariates included age, travel time to the nearest health facility, presence of an older eligible child in the same household, presence of a younger eligible child in the same household, whether the household was between facilities, and which health facility catchment area the household was in. The Gaussian process term accounted for spatial variation driven by unobserved factors and was given a Matern covariance structure parameterised by the range, *ρ*, and marginal variance, *σ*^2^. A household-level random *effect*, 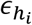, was also included (where *h*_*i*_was the household that child *i* lived in) to account for repeated sampling from households containing multiple eligible children.

The Bayesian model was completed by placing priors of the model parameters. Normal priors were placed on *β*_0_ and *β* with mean 0 and standard deviation 1. Penalised complexity priors were used for the Mátern covariance parameters [29].

To map predicted zero-dose prevalence across the study area, each model was re-fit with only spatial covariates included (i.e., travel time to the nearest health facility and whether the location was between facilities).

#### Effectiveness of additional outreach vaccination sites

The mass vaccination campaign was conducted at health facilities and temporary outreach vaccination sites set up across Choma District. We evaluated the effect of additional outreach vaccination sites in future campaigns. First a geostatistical model was fit to model the probability of a measles zero-dose child identified in the pre-campaign enumeration being vaccinated during the mass vaccination campaign. This model structure was the same as the zero-dose prevalence model previously described, with a child being vaccinated in the campaign modelled as a Bernouilli process based on an underlying probability. This probability was modelled as the sum of a linear combination of covariates and a Gaussian process term, again as in the zero-dose prevalence model. Here the covariates were age and travel time to the nearest campaign site. Campaign sites were defined as health facilities or outreach vaccination sites. After fitting the model, the probability of a measles zero-dose child of a given age and location being vaccinated in the campaign could be calculated. Let *p*(*a, l*) be this probability for a child of *a* months of age at location *l*.

To evaluate the effect of adding an outreach vaccination site at a given location *s*, travel times to the nearest campaign site were recalculated with this additional site included. The fitted relationship was then used to calculate an updated probability, 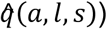, of a measles zero-dose child of a given age at a given location being vaccinated in the campaign. We assumed that individuals would not travel more than 60 minutes to be vaccinated and hence, the final probability of a child being vaccinated given this additional vaccination site was defined to be 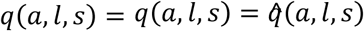 if *l* and *s* were within 60 minutes travel time of each other and *q*(*a, l, s*) = *p*(*a, l, s*) otherwise.

The overall effect of adding this outreach vaccination site, *Eff*(*s*), was defined as the difference between these probabilities, summed over all measles zero-dose children identified in the survey,

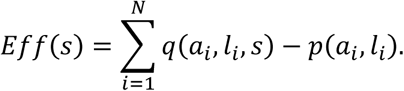

In other words, the total effectiveness of an additional site was defined as the increase in vaccination probability of a child of a given age and location integrated over the empirical age and spatial distribution of measles zero-dose children. The effect of adding multiple additional outreach vaccination sites at different locations was similarly calculated by recalculating the travel times and vaccination probabilities.

An alternative form of the geospatial model for vaccination probability was also fit in which time to the nearest health facility and time to the nearest outreach vaccination site were included as separate covariates to investigate the sensitivity of our results to combining both (as time to the nearest campaign site) in the main analysis.

#### Model validation

Both geostatistical models were validated using k-fold cross-validation. Evaluating model performance by at a household level is challenging due to high sampling variance of the data generating process at low prevalence values and may not be the most relevant metric from an operational perspective, as even targeted activities are unlikely to be planned at this level. A more relevant spatial scale is performance for small clusters of households, which we term settlements, that are used for planning of other public health measures in Zambia such as indoor residual spraying for malaria control [30,31]. Observed and predicted prevalence was compared at the settlement level, where these settlements were groups of households generated by k-means spatial clustering, intended to approximate the true settlements in the study area. The number of households per cluster and the number of folds for k-fold cross-validation was varied (see Supplementary information section 4.1). We also investigated the potential for using the zero-dose prevalence model to predict prevalence in new locations by fitting the model on data from 3 catchment areas and making predictions for all other catchments.

An assumption underlying our analysis of the effect of adding outreach vaccination sites in different locations was that the learned relationship between travel time to the nearest campaign site and probability of a measles zero-dose child being vaccinated in the campaign reflected the causal effect of campaign sites. If this relationship was confounded, however, this may not necessarily be the case. For example, if some unobserved factors (such as accessibility) caused outreach vaccination sites to be located in areas where children were already more likely to be reached by the campaign, this could inflate the apparent effect of outreach vaccination sites. A negative control [32] was used to check whether such confounding existed. A negative control is a response variable that is similar to the response variable of interest (in this case whether a measles zero-dose child was vaccinated during the campaign) but which is known or believed to be unaffected by the explanatory variable (in this case distance to the nearest campaign site). The analysis is then repeated with this alternative response variable and if the learned relationship with the explanatory variable is non-zero then it suggests this analysis is confounded, and therefore the main analysis may also be confounded. We used measles zero-dose status before the campaign, which clearly could not be affected by the campaign itself, as a negative control to investigate any potential confounding.

## RESULTS

In total, 41,952 structures were identified within the study area from aerial satellite imagery. Of these, 10,758 households were eligible for the study (with many households consisting of multiple structures). In the pre-campaign enumeration phase, 13,519 children were registered and eligible for the study, of whom 1,870 (13.8%) were younger than 9 months and 11,649 (86.2%) were 9-60 months old (Figure 2). Of the children younger than 9 months, 322 (17.3%) had not received DTP1 and were therefore classified as DTP zero-dose children. Four hundred and seventy (4.3%) children 9-60 months of age had not received MCV1 and were classified as measles zero-dose.

**Figure 2:**
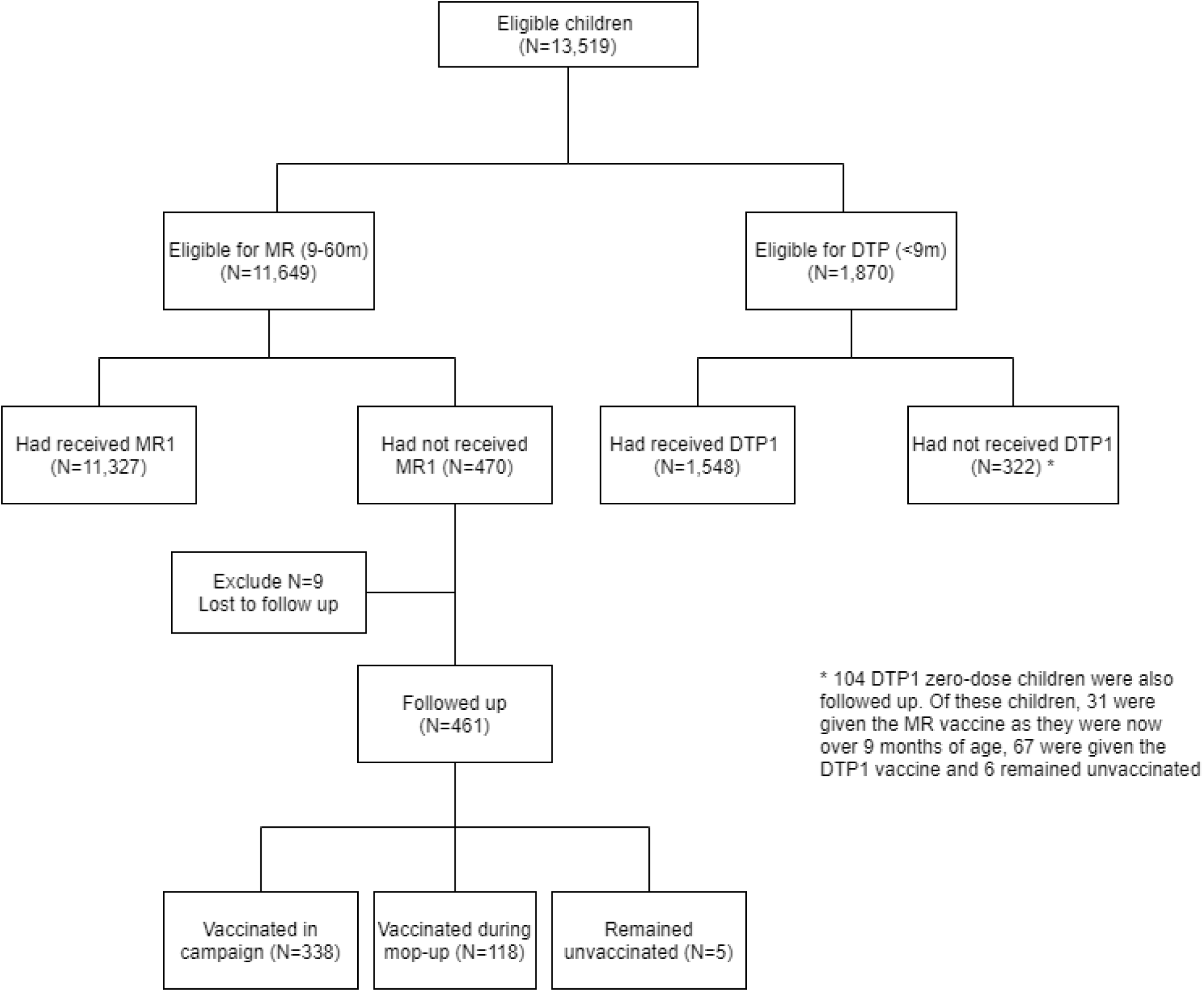
Flow chart describing the data collected.

Approximately one week after the mass measles and rubella vaccination campaign, 461 of the measles zero-dose children identified in the pre-campaign enumeration were successfully followed up as part of a targeted mop up activity. Of these, 338 (73.3%) received the measles and rubella vaccine during the campaign, 118 (25.6%) were vaccinated during this mop up activity, and 5 (1.1%) remained unvaccinated. During these mop up activities, some of DTP zero-dose children identified in the initial survey were also followed-up and some additional children were registered, although this was not the focus of the CHVs and was carried out in a non-systematic way, detailed in Supplementary material section 1.

There was substantial heterogeneity in DTP and measles zero-dose prevalence prior to the mass vaccination campaign at the health facility catchment-level (Table 1). The lowest prevalence for both was in Batoka (DTP1 = 7.1% and MCV1 = 0.2%), with the highest DTP zero-dose prevalence in Kamwanu (58.9%) and measles zero-dose prevalence in Mapanza (8.2%). There were no clear long-range spatial trends (e.g., north to south) in DTP or measles zero-dose prevalence across the study area. At the sub-catchment level there was some evidence of spatial clustering only in Shampande and Mapanza catchment areas (Moran’s I, p < 0.01, supplementary material section 5).

**Table 1:** Observed DTP and measles zero-dose prevalence before the measles and rubella vaccination campaign by catchment area.

| Catchment | DTP zero-dose prevalence | Measles zero-dose prevalence |
| --- | --- | --- |
| Batoka | 7.1% | 0.2% |
| Choma Railway Surgery | 18.6% | 4.7% |
| Kamwanu | 58.9% | 3.7% |
| Macha | 15.6% | 3.7% |
| Mangunza | 15.2% | 3.6% |
| Mapanza | 14.3% | 8.2% |
| Masuku Mission | 21.7% | 7.2% |
| Mochipapa | 21.3% | 5.7% |
| Nalituba | 18.8% | 2.4% |
| Shampande | 14.8% | 3.1% |

Both DTP and measles zero-dose prevalence prior to the mass vaccination campaign decreased as age increased, and initially increased as travel time to the nearest health facility increased (Figure 3a, 3c). As travel time increased further, DPT zero-dose prevalence increased but measles zero-dose prevalence plateaued, although in both cases there was substantial uncertainty at these distances due to small numbers of observations. DTP and measles zero-dose prevalence was higher in children living in a household between facilities or with other eligible children, with both effects stronger for measles zero-dose status. The proportion of measles zero-dose children vaccinated during the mass vaccination campaign decreased as travel time to the nearest vaccination site increased (Figure 3d).

**Figure 3:**
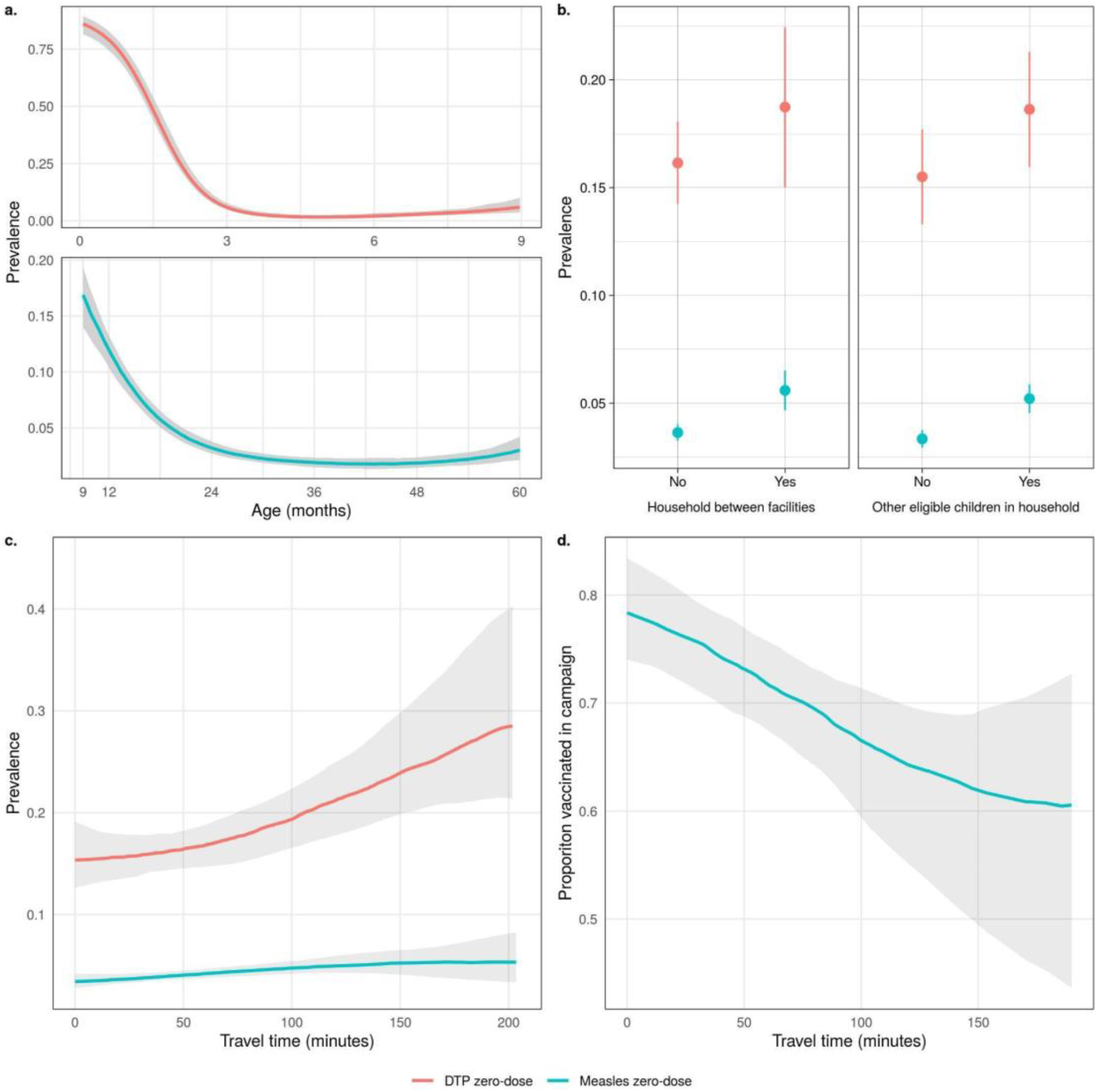
Univariate relationships between DTP and measles zero-dose prevalence and different factors with 95% credible intervals. For continuous variables (a, c, d) a non-parametric model was fit (see Supplementary material section 7). (a) DTP and measles zero-dose prevalence before vaccination campaign by age, (b) DTP and measles zero-dose prevalence before vaccination campaign by whether a child lived in a household between facilities or containing other eligible children, (c) zero-dose and measles zero-dose prevalence before vaccination campaign by travel time to the nearest health facility, and (d) probability of a measles zero-dose child being vaccinated during the campaign by travel time to the nearest campaign site.

When the joint relationship between these factors and DTP zero-dose prevalence was investigated using the geostatistical model, we found broadly similar results to the univariate analyses. An increase in age and the presence of another younger eligible child in the household were associated with increased DTP zero-dose prevalence (Table 2) The effects of the other covariates were uncertain, with the 95% credible intervals containing zero (Table 2). The magnitude of coefficient for age was particularly large, reflecting the initial steep decline in DTP zero-dose prevalence with age (Figure 3). There was no clear association between most health facility catchment areas and DTP zero-dose prevalence except for Kamwanu and Nalituba, which were associated with increased and decreased prevalence, respectively.

**Table 2:** Mean and 95% credible interval for covariate effects in the DTP and measles zero-dose prevalence models.

| Covariate | Mean (CI) |  |
| --- | --- | --- |
|  | DTP zero-dose | Measles zero-dose |
| Time to nearest facility | 0.12 (-0.21, 0.45) | 0.14 (0.01, 0.28) |
| Between facilities | -0.08 (-0.58, 0.41) | 0.21 (-0.05, 0.46) |
| Age | -3.31 (-3.7, -2.91) | -0.83 (-0.96, -0.71) |
| Other older child in household | -0.07 (-0.43, 0.29) | 0.3 (0.08, 0.51) |
| Other younger child in household | 0.99 (0.01, 1.98) | 0.15 (-0.12, 0.41) |
| Urban | -0.23 (-1.61, 1.16) | 0.17 (-1.06, 1.4) |
| Batoka | 0.97 (-0.96, 2.9) | -2.17 (-3.2, -1.13) |
| Choma Railway Surgery | 0.26 (-1.01, 1.52) | 0.28 (-0.89, 1.45) |
| Kamwanu | 4.70 (2.98, 6.41) | -0.03 (-0.81, 0.76) |
| Macha | -1.05 (-2.21, 0.11) | 0.07 (-0.68, 0.82) |
| Mangunza | -0.93 (-2.13, 0.27) | 0.06 (-0.69, 0.82) |
| Mapanza | -0.84 (-2.2, 0.51) | 1.02 (0.29, 1.75) |
| Masuku Mission | -0.89 (-2.12, 0.35) | 0.73 (0.00, 1.47) |
| Mochipapa | -0.47 (-1.71, 0.77) | 0.53 (-0.22, 1.28) |
| Nalituba | -1.26 (-2.52, 0.00) | -0.39 (-1.29, 0.5) |
| Shampande | -0.49 (-1.73, 0.76) | -0.11 (-1.28, 1.05) |

The results of the geostatistical model of measles zero-dose prevalence were also similar to the univariate analyses. Measles zero-dose prevalence decreased with age (at a slower rate than DTP zero-dose prevalence), while an increase in travel time to the nearest facility and the presence of an older eligible child in the household were both associated with increased measles zero-dose prevalence. The posterior mean for the effect of a household being between facilities was also positive, although the 95% credible interval for this coefficient narrowly contained zero. Batoka catchment was associated with lower measles zero-dose prevalence while Masuku Mission and Mapanza catchments were associated with increased prevalence.

Many of the patterns in the maps of predicted zero-dose prevalence (Figures 4a and 4b) reflect the catchment-level patterns in the observed data, such as substantially higher DTP zero-dose prevalence in Kamwanu and lower measles zero-dose prevalence in Batoka. There is also subdistrict heterogeneity apparent in both maps, driven by the spatial covariates relating to health facility access and trends in the observed data learned by the Gaussian process term in the model. The model performed well under 10-fold cross validation (Figure 4c), with correlations between the observed and predicted prevalence at the settlement level of 0.632 and 0.534 for children younger than 9 months and 9 months and older respectively (see supplementary material section 4 for details of additional validation).

**Figure 4:**
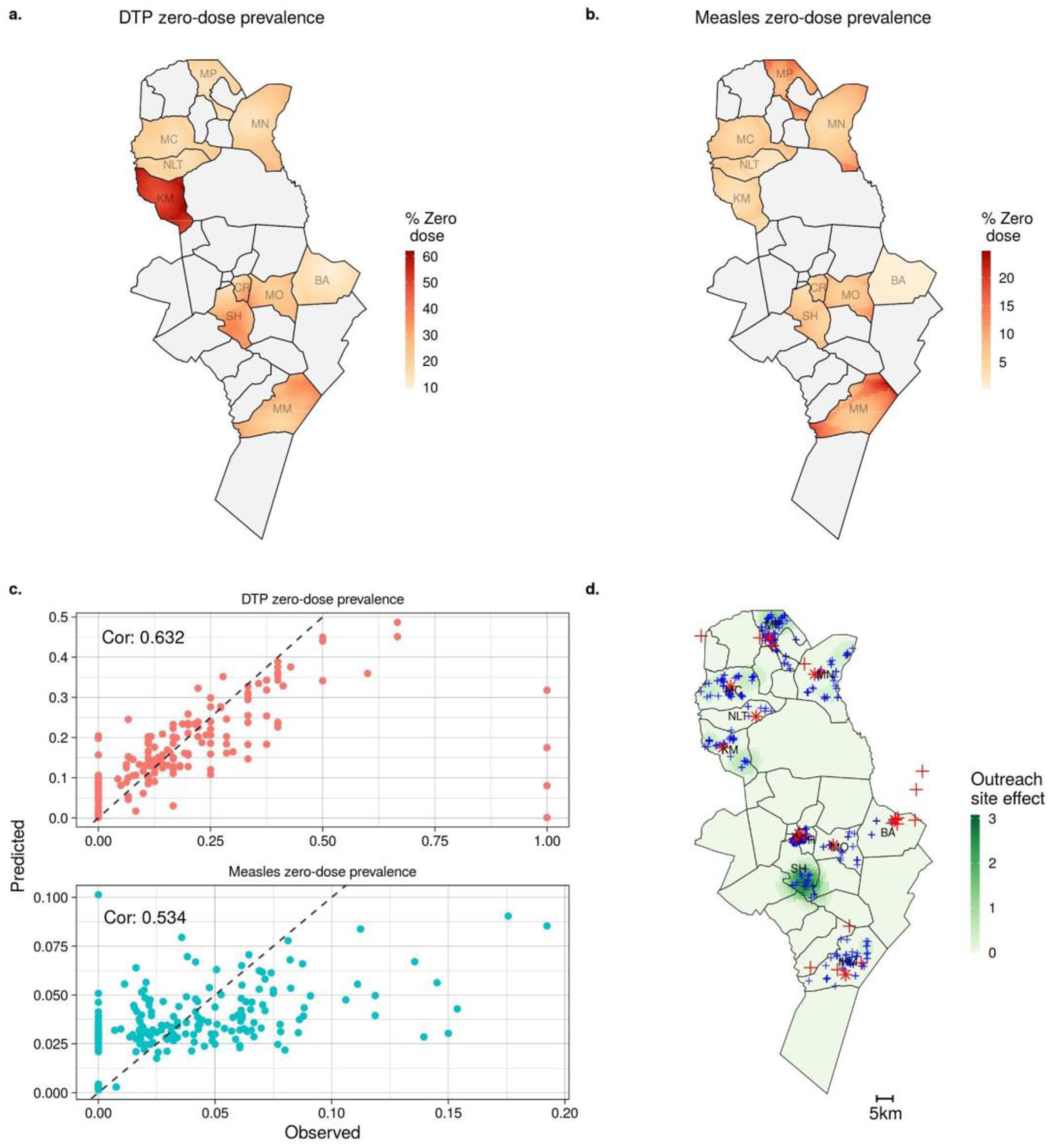
Results from geostatistical models. (a) and (b) Predicted DTP and measles zero-dose prevalence before the mass vaccination campaign respectively. (c) Predicted and observed DTP and measles zero-dose prevalence during cross-validation at the settlement level. (d) Predicted effectiveness (overall increase in vaccination probability over all measles zero-dose children) of adding an additional vaccination site in each location (shown in green with darker green representing greater effectiveness), with locations of measles zero-dose children shown by blue crosses, health facilities as red stars and current outreach sites as red crosses.

Table 3 shows the fitted coefficient values from the geostatistical model of the probability of a measles zero-dose child being vaccinated during the mass vaccination campaign. Increased distance from a campaign site was associated with decreased probability of vaccination. There was no clear relationship between vaccination during the mass vaccination campaign and age. Magunza and Mochipapa catchment areas were associated with increased probability of vaccination while Macha catchment was associated with decreased probability. When the alternative form of the model was fit, in which time to the nearest health facility and time to the nearest outreach vaccination site were considered as separate covariates, there was no significant difference in results (see supplementary material section 4.3).

**Table 3:** Mean and 95% credible interval for covariate effects in the model of probability of a measles zero-dose child being vaccinated during the mass measles and rubella vaccination campaign.

| Covariate | Mean (CI) |
| --- | --- |
| Time to nearest campaign site | -0.52 (-0.74, -0.30) |
| Age | -0.10 (-0.31, 0.11) |
| Batoka | 0.32 (-1.10, 1.75) |
| Choma Railway Surgery | -0.77 (-1.63, 0.09) |
| Kamwanu | -0.72 (-1.61, 0.17) |
| Macha | -1.05 (-1.87, -0.23) |
| Mapanza | -0.17 (-0.97, 0.63) |
| Mangunza | 1.02 (0.01, 2.02) |
| Masuku Mission | -0.10 (-0.99, 0.79) |
| Mochipapa | 1.58 (0.26, 2.90) |
| Nalituba | -0.72 (-1.95, 0.52) |
| Shampande | 0.61 (-0.21, 1.42) |

The results of the geostatistical model of vaccination probability were used to estimate the effect of placing additional vaccination sites in different locations (these results are shown in Figure 4d). The greatest estimated impact was in the south of Shampande catchment, in the center of Choma District, where there were many measles zero-dose children with no campaign site within a 60 minute walk. Another area with relatively high impact is in the north of Mapanza catchment, in the north of the district, where again there were measles zero-dose children relatively far from any existing campaign site. There were no areas in the Choma Railway Surgery, Mochipapa and Batoka catchment areas that would have benefited from additional vaccination sites, likely due to already high vaccination probability and low measles zero-dose prevalence in Batoka and Mochipapa or high vaccination site coverage in Choma Railway Surgery.

The optimal locations for three additional sites are shown in Supplementary Figure S7 and follow a similar pattern to optimizing the effect of a single site, with additional sites in Shampande, Mapanza and Kamwanu. The alternative analysis using measles zero-dose status as negative control found no evidence of confounding (see supplementary material section 4.4).

## DISCUSSION

Despite substantial progress in expanding vaccination coverage, there remain zero-dose children and missed communities that are at risk of outbreaks, stalling progress towards disease control and elimination. In areas with high routine vaccination coverage, identifying these communities is necessary to improve targeted and tailored outreach vaccination services. The results of this work highlight substantial fine-scale spatial heterogeneity in the prevalence of zero-dose children in rural Zambia. For both DTP and measles, there was significant variability in vaccination coverage between health facility catchment areas, some evidence of sub-catchment clustering of zero-dose children, and variation in coverage based on access to healthcare. There was also evidence of fine-scale spatial variation in the effectiveness of the mass measles and rubella vaccination campaign in reaching measles zero-dose children.

This heterogeneity has important implications for disease control, as areas with a high proportion of undervaccinated children may be at risk of outbreaks despite high coverage overall (over 95% of children 9-60 months in the study had received MCV1 before the mass vaccination campaign). The observed increase in zero-dose prevalence as travel time to the nearest health facility increased suggests that travelling long distances may be an impediment to vaccination, as has been previously observed [14,18,19]. This is also supported by the decrease in proportion of measles zero-dose children vaccinated during the mass vaccination campaign as distance to the nearest vaccination increased. There was also an apparent increase in zero-dose prevalence in areas approximately equidistant from two health facilities and this effect remained for children 9-60 months when controlling for other factors, including travel time to the nearest facility. This may indicate that households in these areas are not being served by either of the nearest health facilities. The high rates of vaccination during the mop-up activities of measles zero-dose children not reached by the campaign suggests that inadequate access to healthcare or prioritization are more likely causes of undervaccination in the study area than vaccine refusal.

The presence of another eligible child in the household was also associated with increased likelihood of DTP and measles zero-dose status, a similar result to the lower levels of childhood vaccination in larger families found in some studies [19,33,34]. The presence of another child in the household could affect vaccination status directly or this result may reflect correlations between household size and other socioeconomic factors that influence vaccination rates.

Recently the Gavi, global funding agencies, and some national vaccination program managers have advocated targeted and tailored measles and rubella vaccination activities as opposed to non-selective, nationwide campaigns [35]. To target intensified periodic routine immunization activities, it is important to understand the spatial distribution of the zero-dose children or missed communities at a finer, subnational scale. Our analysis of the effectiveness of additional outreach vaccination sites shows how fine-scale spatial data can be used to answer operationally important questions and target allocation of vaccination resources to communities with where there are likely to be more zero-dose children. Several areas were identified that had high numbers of measles zero-dose children and were relatively far from existing campaign sites. If outreach vaccination sites were set up in these areas, there is an increased likelihood that measles zero-dose children would be reached. Moreover, by quantifying the effectiveness of the measles and rubella vaccination campaign in reaching measles zero-dose children, we were able to compare the relative impact of adding one or more vaccination sites in different locations. While real-world decision-making regarding planning and targeting of future campaigns will depend on many additional factors, analyses such as these could be a useful starting point for micro planning of routine and campaign-based vaccination activities.

This work also has implications for the distribution of other vaccines through mass campaigns and for child health programs more generally. Identifying and successfully providing healthcare to individuals with limited access to or interaction with routine health systems is vital for achieving consistently high coverage of public health interventions. These individuals or communities may be found in areas with high proportions of zero-dose children. Furthermore, other child survival interventions, including child health weeks, use much of the same infrastructure and healthcare providers as the mass measles and rubella vaccination campaign. Therefore, zero-dose children who were not vaccinated during the campaign may also be missed by these interventions. Thus, our approach is applicable to other child health interventions and campaigns.

The household survey strategy described here provided a unique source of information on the fine-scale spatial variation in the prevalence of zero-dose children. The process of identifying structures via satellite imagery enabled data collectors to visit households that were not previously known to the survey team and ensured that remote locations were included. In contrast, routinely collected vaccination coverage data are collected at a more aggregated spatial scale, such as at the health facility or district-level, and are therefore likely to mask some of the fine-scale variation. Furthermore, the collection of routine data is likely to be biased towards individuals or communities with better access to the healthcare system and therefore may overestimate coverage. This can be seen in the high coverage values in many districts (often above 100%) where true coverage is believed to be much lower [23,36]. Another common source of vaccination coverage data is from Demographic and Health Surveys (DHS) [37]; however, these data are unable to provide the same level of spatial granularity as the data analyzed here due to privacy-preserving spatial displacement and less dense sampling schemes over much wider areas. In the 2018 DHS in Zambia, for example, there were only four clusters in our study area and eight in Choma District [22].

One limitation of this study is the limited demographic information collected. While the number of questions on the survey was intentionally limited to reduce the time taken to conduct each survey, more demographic information (such as indicators of socioeconomic status) would have allowed more factors potentially influencing vaccination status to be considered. This could have provided more insight into areas likely to have high numbers of zero-dose children in different districts or in the health facility catchment areas not included in the study. Similarly, conducting the survey in more than one district could have improved the generalizability of these findings and informed the reliability of some of the associations found. We performed several model validation steps and found that the model performed well during testing. However, the model may not perform as well in areas with substantially different geographic, and demographic characteristics than Choma District. Further, we focused our analysis on children identified as DTP or measles zero-dose, defined as those that had not received DTP1 or MCV1, respectively. This did not include children who were partially immunized, i.e., those who received MCV1 but not MCV2, who may represent another partially susceptible population. Additionally, data were not collected on DTP1 vaccination status for children 9 months and older, which would have allowed us to compare access to routine services more broadly.

## CONCLUSION

We provide one of the first spatial analyses of the prevalence of zero-dose children and possible strategies to improve targeted and tailored measles and rubella vaccination activities. Our analysis indicates that greater travel time to healthcare (either health facilities or vaccination sites) was associated with increased zero-dose prevalence and decreased likelihood of a measles zero-dose child being vaccinated in the mass vaccination campaign. The presence of other children in the household and living approximately equidistant from two health facilities were also associated with greater zero-dose prevalence. The fine-scale spatial variation in zero-dose prevalence highlights the potential benefits of sub-district targeting and micro-planning of vaccination activities and child health services more broadly.

## Data Availability

Restrictions apply to the availability of these data. Data was obtained under data sharing agreements from Zambia Ministry of Health and the Zambia National Health Research Authority and can only be shared with their permission from the Ministry of Health.

## ACKNOWLEDGEMENTS

This work was only possible through the contributions of many other individuals and organizations. We thank Macha Research Trust organization and all individuals involved in the field activities (Hope Simpito, Wesley Chisenga, Dr. Namani Monze, the health center in charges, environmental health technicians and community health volunteers) for collecting structure and individual data. We thank Akros organization for providing mapping support. We thank the Zambia Ministry of Health for providing guidance on the objectives and for facilitating introductions to the many health facilities. Finally, we are greatly indebted to the Bill and Melinda Gates Foundation for providing us with funding and strategic advice on all aspects of the methods, presentation, and interpretation throughout the project.

## FOOTNOTES

### Contributors

Conceptualization: R.A., S.M., A.W., Y.Y.; data collection: F.D.M., G.M., P.N., K.H., J.A.D.; methodology: R.A., S.M., A.W., Y.Y. ; software and analysis: R.A., Y.Y., K.H., J.A.D. ; writing—original draft preparation: R.A., Y.Y.; writing—review and editing, S.M., A.W., K.H., J.A.D.; visualization, R.A., Y.Y.; supervision, S.M., A.W.; project administration, Y.Y., G.M., K.H.; funding acquisition: S.M. All other authors critically reviewed results. All authors have read and agreed to the published version of the manuscript.

### Patient and public involvement

Patients and/or the public were not involved in the design, or conduct, or reporting, or dissemination plans of this research

### Patient consent for publication

Not required.

## REFERENCE

1. GBD 2020, Vaccine Coverage Collaborators. Measuring routine childhood vaccination coverage in 204 countries and territories, 1980–2019: a systematic analysis for the Global Burden of Disease Study 2020, Release 1. Lancet. 2021 Aug 7;398(10299):503–21.

2. WHO. WHO/UNICEF estimates of national immunization coverage [Internet]. [cited 2021 Aug 25]. Available from: https://www.who.int/teams/control-of-neglected-tropical-diseases/yaws/diagnosis-and-treatment/immunization-analysis-and-insights

3. Causey K, Fullman N, Sorensen RJD, et al. Estimating global and regional disruptions to routine childhood vaccine coverage during the COVID-19 pandemic in 2020: a modelling study. Lancet. 2021 Aug 7;398(10299):522–34.

4. UNICEF. Vaccination and Immunization Statistics [Internet]. UNICEF DATA. 2021 [cited 2021 Aug 25]. Available from: https://data.unicef.org/topic/child-health/immunization/

5. Cadena J, Marathe A, Vullikanti A. Critical Spatial Clusters for Vaccine Preventable Diseases. In: Thomson R, Bisgin H, Dancy C, Hyder A, Hussain M, editors. Social, Cultural, and Behavioral Modeling [Internet]. Cham: Springer International Publishing; 2020 [cited 2021 Aug 25]. p. 213–23. (Lecture Notes in Computer Science; vol. 12268). Available from: https://link.springer.com/10.1007/978-3-030-61255-9_21

6. Ferrari MJ, Bansal S, Meyers LA, et al. Network frailty and the geometry of herd immunity. Proc Biol Sci. 2006 Nov 7;273(1602):2743–8.

7. Anderson RM, May RM. Vaccination and herd immunity to infectious diseases. Nature. 1985 Nov;318(6044):323–9.

8. Chard AN. Routine Vaccination Coverage — Worldwide, 2019. MMWR Morb Mortal Wkly Rep. 2020 Nov 13;69(45):1706–10.

9. WHO, UNICEF. Progress and Challenges with Achieving Universal Immunization Coverage [Internet]. Geneva, Swizerlands; 2020 Jun [cited 2021 Aug 25] p. 1–25. (1). Report No.: 1. Available from: https://www.who.int/publications/m/item/progress-and-challenges-with-achievinguniversal-immunization-coverage

10. WHO. Immunization agenda 2030 [Internet]. Geneva, Swizerlands; 2020 Apr [cited 2021 Aug 25] p. 1–60. Available from: https://www.who.int/publications/m/item/immunization-agenda-2030-a-global-strategy-to-leave-no-one-behind

11. Gavi. Annual Progress Report 2019. Geneva, Swizerlands: Gavi; 2019 Dec p. 1–68.

12. Grundy J, Biggs B-A. The Impact of Conflict on Immunisation Coverage in 16 Countries. Int J Health Policy Manag. 2018 Dec 30;8(4):211–21.

13. Cata-Preta BO, Santos TM, Mengistu T, et al. Zero-dose children and the immunisation cascade: Understanding immunisation pathways in low and middle-income countries. Vaccine. 2021 Jul 22;39(32):4564–70.

14. Cutts FT, Danovaro-Holliday MC, Rhoda DA. Challenges in measuring supplemental immunization activity coverage among measles zero-dose children. Vaccine. 2021 Mar 1;39(9):1359–63.

15. Jani JV, De Schacht C, Jani IV, et al. Risk factors for incomplete vaccination and missed opportunity for immunization in rural Mozambique. BMC Public Health. 2008 May 16;8:161.

16. Abdulraheem IS, Onajole AT, Jimoh AAG, et al. Reasons for incomplete vaccination and factors for missed opportunities among rural Nigerian children. Journal of Public Health and Epidemiology. 2011 Apr;3(4):194–203.

17. Konstantyner T, Taddei JA de AC, Rodrigues LC. Risk factors for incomplete vaccination in children less than 18 months of age attending the nurseries of day-care centres in Sao Paulo, Brazil. Vaccine. 2011 Nov 21;29(50):9298–302.

18. Agócs M, Ismail A, Kamande K, Tabu C, et al. Reasons why children miss vaccinations in Western Kenya; A step in a five-point plan to improve routine immunization. Vaccine. 2021 Aug 9;39(34):4895–902.

19. Rainey JJ, Watkins M, Ryman TK, et al. Reasons related to non-vaccination and under-vaccination of children in low and middle income countries: Findings from a systematic review of the published literature, 1999–2009. Vaccine. 2011 Oct 26;29(46):8215–21.

20. Bosch-Capblanch X, Banerjee K, Burton A. Unvaccinated children in years of increasing coverage: how many and who are they? Evidence from 96 low-and middle-income countries. Tropical Medicine & International Health. 2012;17(6):697–710.

21. Portnoy A, Jit M, Helleringer S, Verguet S. Impact of measles supplementary immunization activities on reaching children missed by routine programs. Vaccine. 2018 Jan 2;36(1):170–8.

22. Zambia Statistics Agency, Ministry of Health, University Teaching Hospital Virology Laboratory, et al. Zambia Demographic and Health Survey 2018 [Internet]. 2020 Jan [cited 2021 Aug 25]. Available from: https://dhsprogram.com/publications/publication-fr361-dhs-final-reports.cfm

23. Ministry of Health Zambia. Annual progress reports-Choma District Health Management Team. Choma, Zambia; 2020 Jan.

24. Reveal. How do you ensure no one is missed by life-saving health services? [Internet]. Reveal. 2021 [cited 2021 Aug 25]. Available from: https://revealprecision.com/

25. Weiss DJ, Nelson A, Gibson HS, et al. A global map of travel time to cities to assess inequalities in accessibility in 2015. Nature. 2018 Jan;553(7688):333–6.

26. Diggle PJ, Tawn JA, Moyeed RA. Model-based geostatistics. Journal of the Royal Statistical Society: Series C (Applied Statistics). 1998;47(3):299–350.

27. Bhatt S, Weiss DJ, Cameron E, et al. The effect of malaria control on Plasmodium falciparum in Africa between 2000 and 2015. Nature. 2015 Oct;526(7572):207–11.

28. Diggle PJ, Giorgi E. Model-Based Geostatistics for Prevalence Mapping in Low-Resource Settings. Journal of the American Statistical Association. 2015 Jun 1;111(515):1096–0020.

29. Fuglstad G-A, Simpson D, Lindgren F, et al. Constructing Priors that Penalize the Complexity of Gaussian Random Fields. 150300256 [stat] [Internet]. 2017 Nov 27 [cited 2021 Aug 25]; Available from: http://arxiv.org/abs/1503.00256

30. Ministry of Health Zambia. National Malaria Elimination Strategic Plan 2017-2021. Lusaka, Zambia: Ministry of Health Zambia; 2017 Jan p. 1–109.

31. Ministry of Health Zambia. Malaria Operational Plan 2020-2021. Lusaka, Zambia: Ministry of Health Zambia; 2020 Jan p. 1–109.

32. Lipsitch M, Tchetgen Tchetgen E, Cohen T. Negative Controls: A Tool for Detecting Confounding and Bias in Observational Studies. Epidemiology. 2010 May;21(3):383–8.

33. Ndiritu M, Cowgill KD, Ismail A, et al. Immunization coverage and risk factors for failure to immunize within the Expanded Programme on Immunization in Kenya after introduction of new Haemophilus influenzae type b and hepatitis b virus antigens. BMC Public Health. 2006 May 17;6(1):132.

34. Shamsul A, Nirmal K, Nazarudin S, et al. Factors Influencing Childhood Immunization Defaulters in Sabah, Malaysia. The International Medical Journal Malaysia. 2012 Jun 1;11(1):17–22.

35. Gavi. Gavi Phase 5 Strategy (2021–2025) [Internet]. 2021 [cited 2021 Sep 8]. Available from: https://www.gavi.org/our-alliance/strategy/phase-5-2021-2025

36. Oliphant NP, Mason JB, Doherty T, et al. The Contribution of Child Health Days to Improving Coverage of Periodic Interventions in Six African Countries. Food Nutr Bull. 2010 Sep 131(3_suppl3):S248–63.

37. USAID. The DHS Program - Quality information to plan, monitor and improve population, health, and nutrition programs [Internet]. DHS Program Demographic and Health Survey. [cited 2021 Aug 25]. Available from: https://dhsprogram.com/

