## Supplementary materials for "Using geospatial models to map zero-dose children: factors associated with zero-dose vaccination status before and after a mass measles and rubella vaccination campaign"

Factors associated with zero-dose vaccination status before and after a mass vaccination campaign and implications for supplementary immunization activities

Rohan Arambepola, Yangyupei Yang, Kyle Hutchinson, Francis Mwansa, Julie Ann Doherty, Frazer Bwalya, Phillimon Ndubani, Gloria Musukwa, William B. Moss, Amy Wesolowski, Simon Mutembo

Supplementary information

1. Additional follow up activities

The focus of the follow up activities was to revisit measles zero-dose children, however some DTP zero-dose children were also followed up with and new children registered during this phase. Of the 322 DTP zero-dose children identified before the campaign, 104 were followed up with. These children were all below 9 months of age during the pre-campaign survey but at the time of the follow up 31 were over 9 months and therefore were eligible for the MR vaccine. Of these children over 9 months at the time of follow up, 25 (80.6%) had been given the MR vaccine during the campaign and the remaining 6 (19.4%) were given the MR vaccine during the mop-up activities. The other 73 zero-dose children followed up with were still below 9 months of age at the time of follow up. Of these children, 67 (91.8%) were given the DTP vaccine during the follow up activities and 6 (8.2%) remained unvaccinated. An additional 191 zero-dose children were identified during the follow up activities, of which 37 were under 9 months and 154 were 9 months or older, who were all vaccinated during the mop-up activities with the relevant vaccine (DTP for under 9 months and MR for 9 months or older).

2. Calculating travel times

The time needed to travel between any two locations was calculated using a friction surface produced by Weiss et al. [17] that divides the study area into 1km^2 pixels and quantifies the time taken to travel (on foot) through any pixel and a least-cost algorithm [SI1] which finds the shortest path between the two locations based on this friction surface. The friction surface is based on a number of factors, including land cover and road networks.

3. Households between facilities

The locations of the households in the study that were classified as between health facilities is shown in Figure S1. For the sensitivity analysis, thresholds of 5%, 10% and 25% were also used to classify between facility households. Figure S2 shows the locations of these households in each case and Figure S3 shows zero-dose prevalence in between facility households and all other households in each case. We can see that there is a consistent pattern of higher zero-dose prevalence in between facility households.

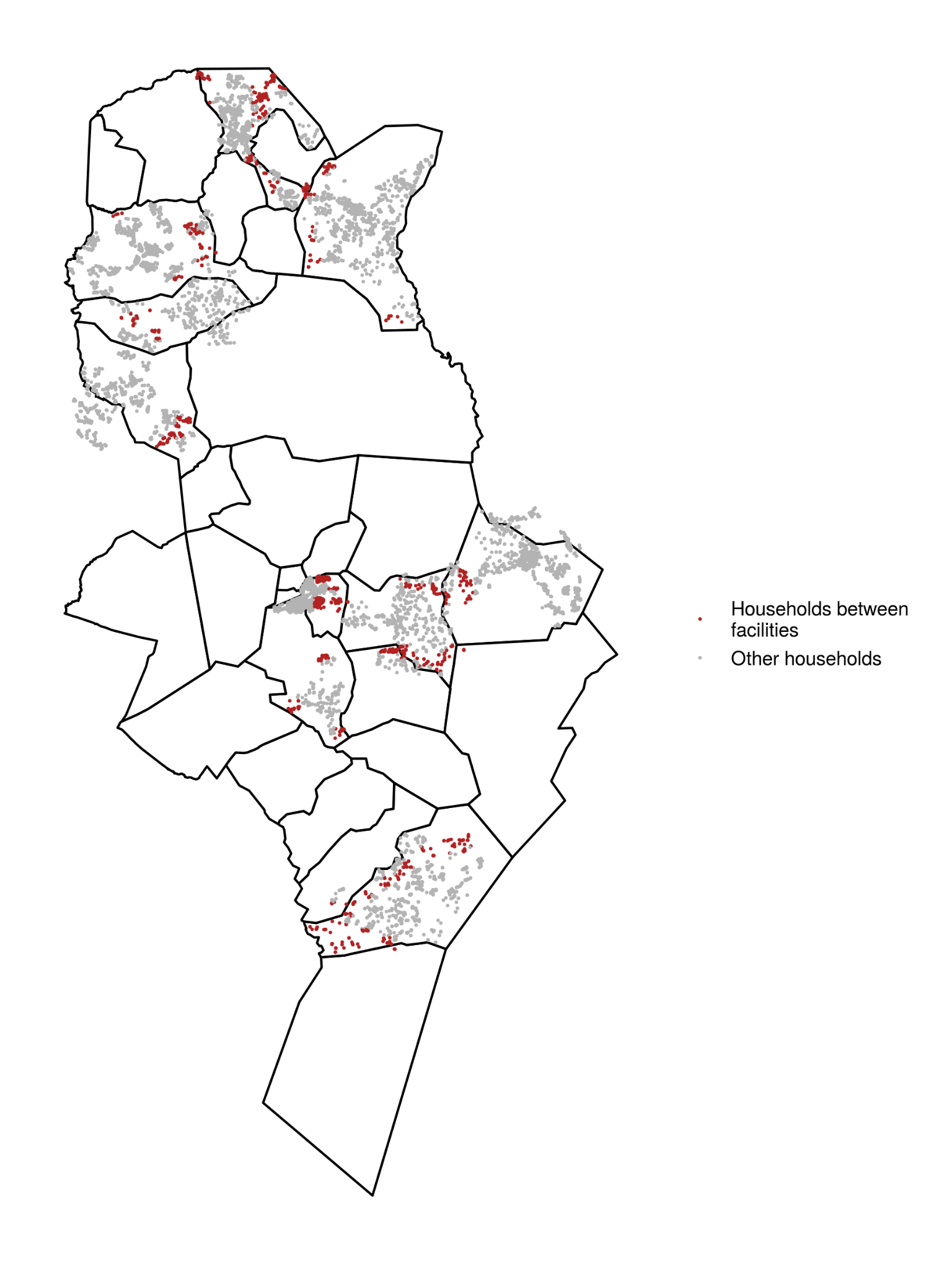

Figure S1: Locations of households in the study shaded based on whether they were classified as between health facilities.

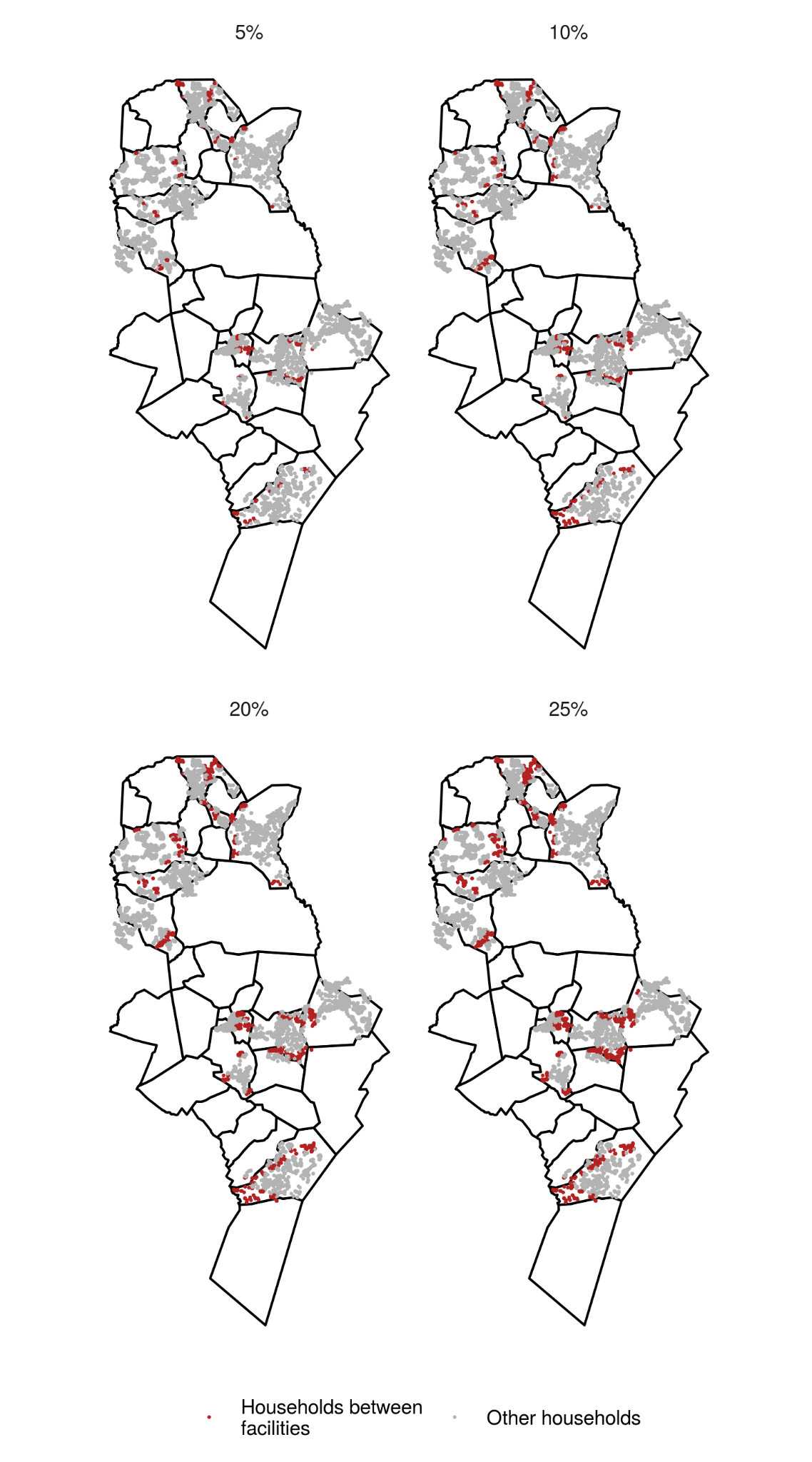

Figure S2: Locations of between facility households with different definition thresholds.

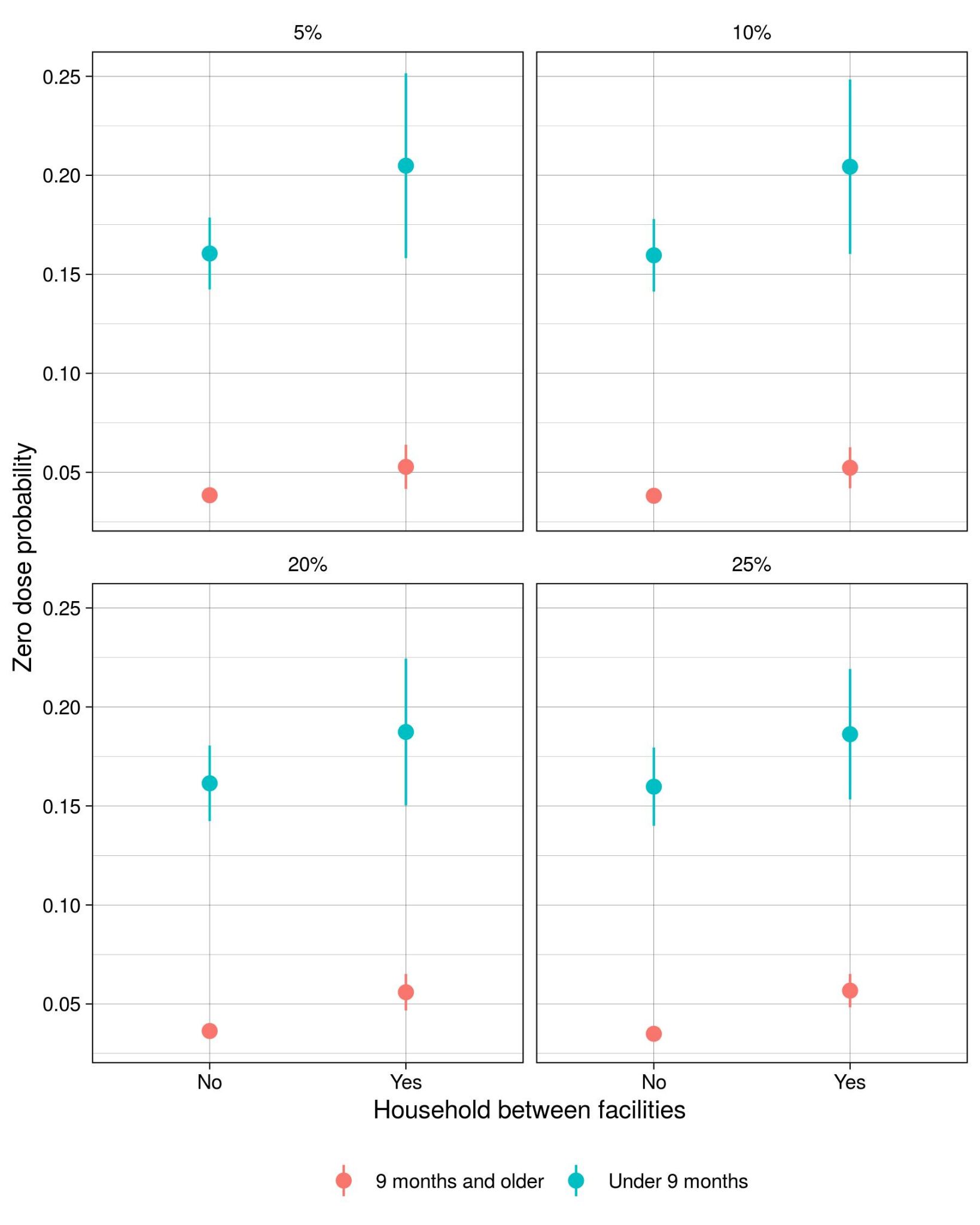

Figure S3: Zero-dose prevalence in between facility households and other households for different thresholds in the definition of a between facility household.

4. Model validation

*4.1 Clustering into settlements and cross-validation:*

Validating the performance of the zero-dose prevalence model at an individual level is challenging due to the relatively high sampling variance (especially for rare events such as zero-dose status, particularly in children 9 months and older). Targeting of vaccination activities is also unlikely to occur at the individual level. A more relevant spatial scale for prediction is the settlement-level, small groups of spatially clustered households. Other public health activities, such as IRS spraying, are already applied at a settlement-level. These settlements are made up of around 250 households on average, although some settlements contain over 1000 households [ref?].

Information on which households in the study belonged to the same settlements was not available, so for the purpose of model validation mock settlements were generated using spatial clustering. The k-means algorithm was used to cluster households based on their locations. The number of clusters (in this case, settlements) in this algorithm is pre-selected. The model was validated using 50, 100 and 200 settlements, corresponding to approximately 215, 108 and 54 households on average per settlement.

K-fold validation was performed by partitioning the settlements into k approximately equal sized folds. The model was then fit k times, with all households in settlements in one fold held out during the fitting process. Each time predictions were then made for the held-out households. Finally, predictions and true values were aggregated to the settlement-level are compared. The correlation between true and observed values for different settlement sizes and numbers of folds is detailed in Tables S2 and S3. Figure S4 shows the households clustered into settlements.

The model of the probability of a zero-dose child being vaccinated during the SIA campaign could not be validated at the settlement level, as the majority of children in each settlement (both the true settlements or those generated by the clustering algorithm) were not zero-dose and therefore were not included in this model. Therefore this model was validated on the individual level, despite the challenges of high sampling variance. On average there was a correlation of 0.37 between the true vaccination status and the predicted probability of vaccination during the campaign when 5- and 10-fold cross-validation was repeatedly performed. The utility of these cross-validated predictions as a binary classifier was also investigated. Figure S5 shows the receiver-operator curve from this analysis. Again, the model performed fairly well, with an area under the curve (AUC) of 0.769.

Table S2: Correlation between true and predicted settlement-level zero-dose prevalence for children under 9 months before the vaccination campaign when using different numbers of settlements and cross-validation folds.

|  | Settlements | | |
| --- | --- | --- | --- |
|  | 50 | 100 | 200 |
| 5 folds | 0.64 | 0.67 | 0.54 |
| 10 folds | 0.65 | 0.68 | 0.53 |

Table S3: Correlation between true and predicted settlement-level zero-dose prevalence for children 9 months and older before the vaccination campaign when using different numbers of settlements and cross-validation folds.

|  | Settlements | | |
| --- | --- | --- | --- |
|  | 50 | 100 | 200 |
| 5 folds | 0.63 | 0.57 | 0.60 |
| 10 folds | 0.71 | 0.62 | 0.63 |

Figure S4: Households clustered into different numbers of settlements. Each point is the location of a household in the study with the colour representing the settlement that households belongs to.

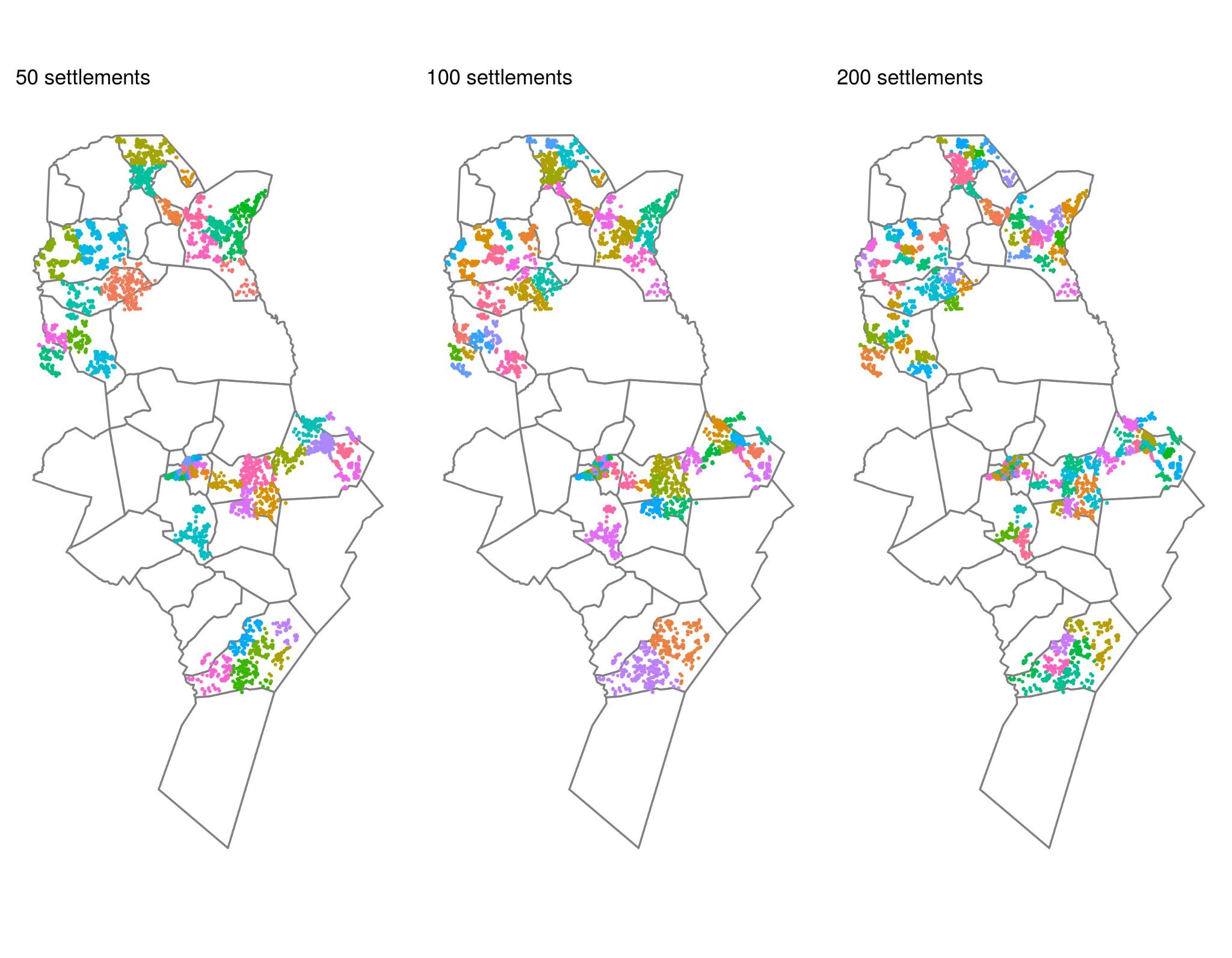

Figure S5: Receiver operator curve for predictions of zero-dose children being vaccinated during the SIA campaign.

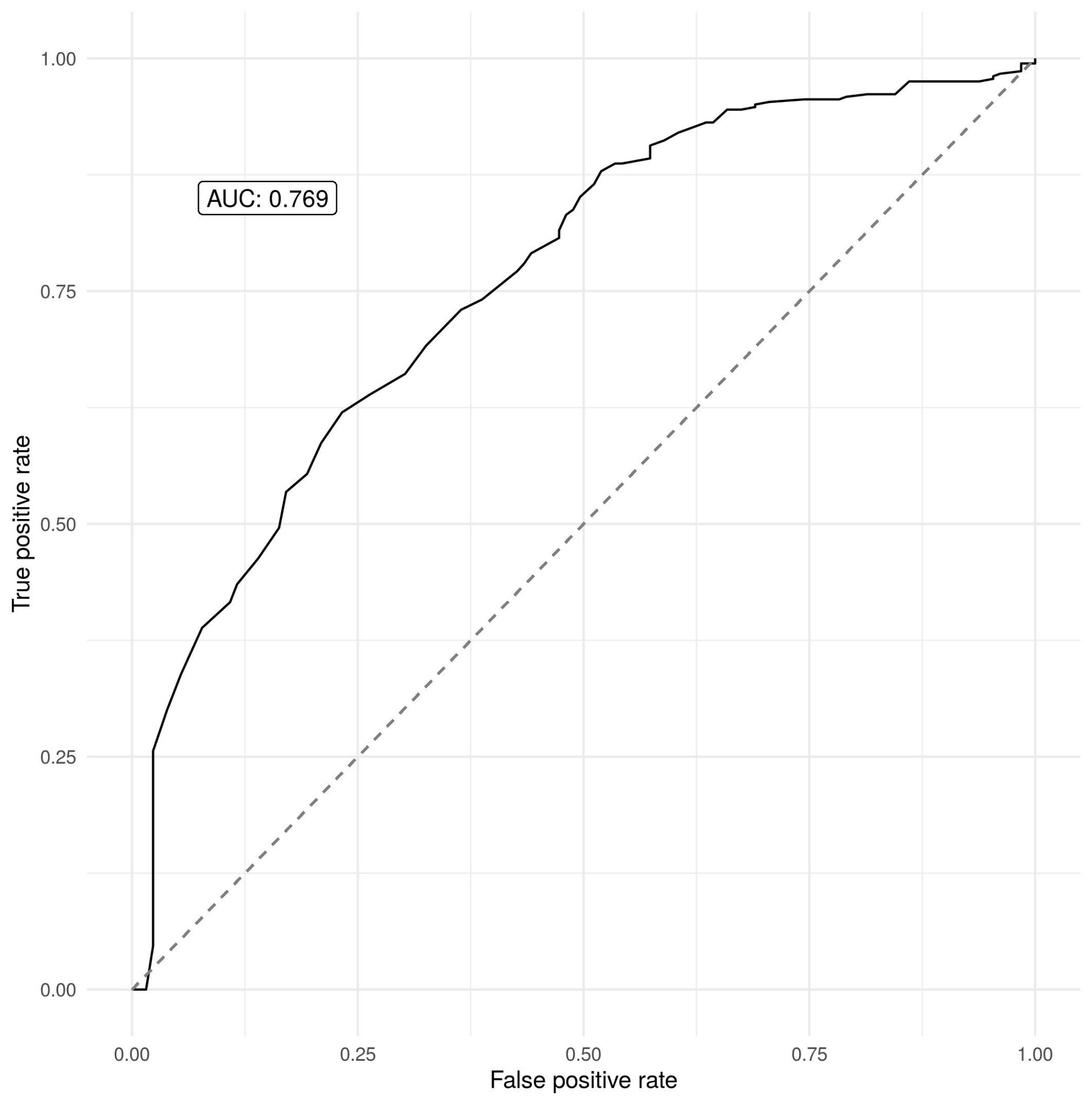

To investigate the ability of the model to predict zero-dose prevalence for completely unobserved catchment areas, three of the catchment areas were left out during the fitting process each time and then prevalence predictions were made for these areas. On average, there was a correlation of 0.348 between predicted and observed DTP zero-dose prevalence and 0.204 for measles zero-dose prevalence. This suggests that some of the variation in zero-dose status in these unobserved catchment areas was successfully predicted but including more explanatory variables (such as socioeconomic variables) could help with this spatial extrapolations.

*4.3 Fitting probability of vaccination model with health facilities and outreach sites considered separately*

As described in the main text, travel time to the nearest vaccination site is included as an explanatory variable in the geostatistical model of the probability of a zero-dose child being vaccinated in the SIA campaign. The fitted relationship is then used to predict the effectiveness of placing additional vaccination sites in different locations. However, if health facilities have a greater effect on vaccination during the campaign (for example, if health facilities have greater capacity to vaccinate or are open for longer) then combining health facilities and outreach sites in this way may overstate the effectiveness of outreach sites. To check this, the model was refit with travel time to the nearest health facility and the nearest outreach site included as separate explanatory variables. The fitted effect of travel time to the nearest outreach site was -0.51 (CI: -0.99, -0.03) which is similar to the effect of travel time to the nearest vaccination site in the original analysis, and is in fact greater in magnitude than the effect of travel time to the nearest health facility which was -0.39 (-0.63, -0.14).

*4.4 Using a negative control to detect confounding*

The relationship learned between the travel time to the nearest vaccination site and probability of vaccination in the campaign by the geospatial model is negative, suggesting that children who lived closer to vaccination sites were more likely to be vaccinated during the campaign. However, using this relationship to predict the effectiveness of adding outreach sites in different locations implicitly assumes that this learned relationship represents the causal effect of outreach sites on vaccination probability. However, if there exist confounding variables (variables that influence both the location of the outreach sites and the likelihood of vaccination) then some or all of the observed relationship could be driven by these factors. For example, if outreach sites were more likely to be placed in less remote areas and zero-dose children in these areas were already more likely to be vaccinated in the campaign, then the observed relationship may be greater than the causal effect of the outreach sites.

A negative control can be used to detect possible confounding by unobserved variables. A negative control is a response variable that is similar to the response variable of interest but that the explanatory variable can have no effect on. If the inferred effect on the explanatory variable on the negative control is non-zero, then this suggests that this effect is due to unobserved confounding and that there might be confounding in the original analysis. In this case our negative control is vaccination status before the vaccination campaign. Clearly the location of outreach sites can have no causal effect on vaccination status before the campaign. However, if the relationship between outreach site location and vaccination in the campaign is confounded by, for example, accessibility then we might expect the relationship between outreach site location and vaccination before the campaign (the negative control) to also be confounded. Figure S6 shows that there is no clear relationship between travel time to the nearest outreach site and vaccination status before the campaign, which therefore gives us more confidence that the learned effect of travel time to the nearest outreach site in the original analysis is not confounded and represents the causal effect. Similarly, Table S4 shows the fitted coefficients of a geostatistical model (with the same design as described in the main text) of measles zero-dose status before the campaign as a function of travel time to the nearest outreach site, travel time to the nearest health facility, age and catchment. Again we see that travel time to the nearest outreach site has no significant effect (95% CI contains zero).

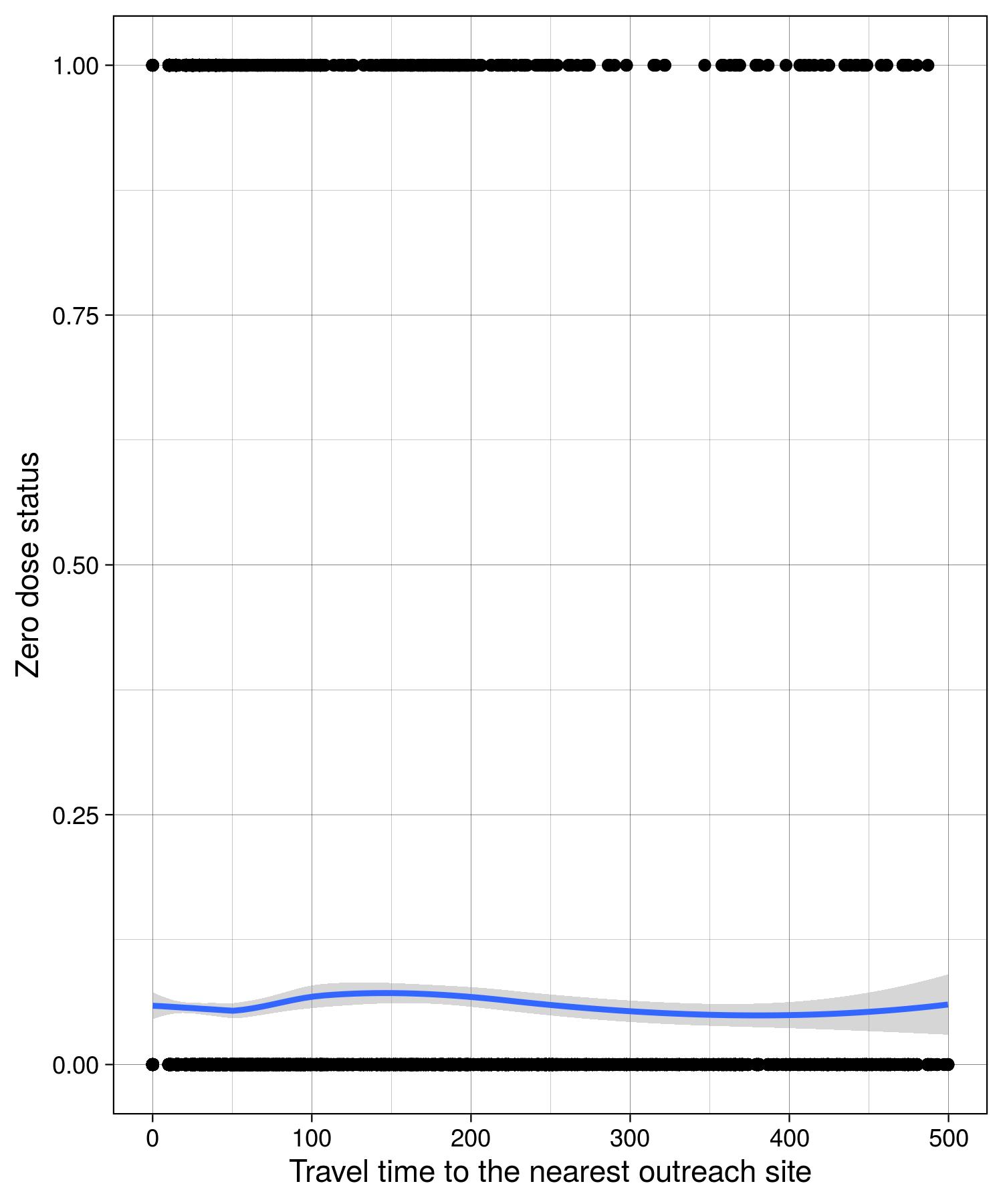

Figure S6: Observed measles zero-dose status before mass vaccination campaign against travel time to the nearest campaign outreach site (in minutes). Observations are shown as black dots and the blue line shows the smoothed relationship.

| Covariate | Mean (CI) |
| --- | --- |
| Time to nearest health facility | 0.14 (0.01, 0.29) |
| Time to nearest outreach site | 0.19 (-0.13, 0.53) |
| Age | -0.10 (-0.31, 0.11) |
| Shampande | 0.10 (-0.60, 0.80) |
| Masuku Mission | 0.82 (0.07, 1.56) |
| Nalituba | -0.63 (-1.55, 0.29) |
| Mochipapa | 0.52 (-0.18, 1.23) |
| Choma Railway Surgery | 0.67 (-0.05, 1.39) |
| Kamwanu | -0.56 (-1.55, 0.43) |
| Batoka | -2.17 (-3.19, -1.14) |
| Macha | -0.08 (-0.82, 0.65) |
| Mangunza | 0.16 (-0.58, 0.90) |
| Mapanza | 1.16 (0.46, 1.87) |

Table S4: Fitted coefficients from geostatistical model of measles zero-dose status before mass vaccination campaign

5. Testing for sub-catchment clustering

Moran’s I [SI2] was used to test for clustering of zero-dose status for each health facility catchment using an exponential weight function. Shampande and Mapanza catchments had p-values less than 0.01.

6. Optimising for multiple additional vaccination outreach sites

Multiple additional vaccination sites were optimised sequentially: First, the optimal location for one additional vaccination site was found. Next, travel times and vaccination probabilities were updated under the assumption that an outreach site had been added at this location. Then the optimal location of another vaccination site was found. This was repeated for three sites, though in theory could be continued indefinitely. The same objective function could also be used to optimise the addition of multiple locations jointly.

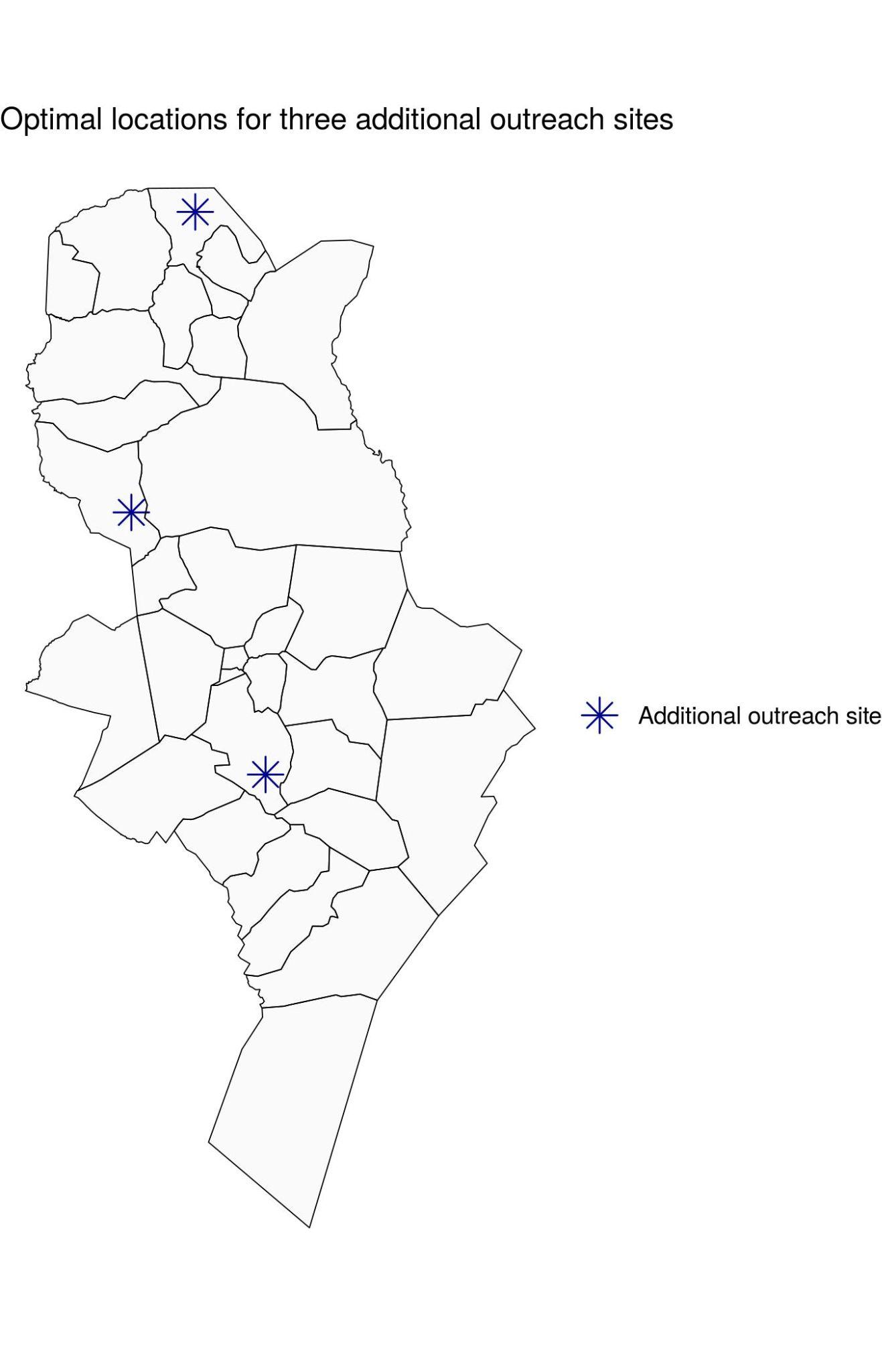

Figure S7: Optimal locations for three additional outreach vaccination sites

7. Non-parametric model for investigating univariate relationships

A non-parametric model was used to investigate the univariate relationship between continuous explanatory variables (age, travel time to the nearest health facility, travel time to the nearest campaign site) and vaccination status. Let $y_{i}$ be the vaccination status for observation i (i.e. 1 if vaccinated and 0 if not). This was modelled as a bernouilli random variable with underlying probability $p_{i}$. This probability was modelled on a logit scale as the sum of a constant and a Gaussian process over the explanatory variable, i.e.

$logit(p_{i}) = \beta_{0}+ f(x_{i})$

where $f$ was the Gaussian process term, $x_{i}$ was the value of the explanatory variable for this observation (e.g. age) and $\beta_{0}$ was a constant. The constant and Gaussian process term were learned during the fitting process. An exponential kernel function was used for the Gaussian process.

References

[SI 1] Dijkstra, Edsger W. "A note on two problems in connexion with graphs." *Numerische mathematik* 1.1 (1959): 269-271.

[SI 2] Moran, Patrick AP. "Notes on continuous stochastic phenomena." Biometrika 37.1/2 (1950): 17-23.
